# Anti-Nucleocapsid and Anti-Spike Antibody Trajectories in People with Post-Covid Condition versus Acute-Only Infections: Results from the Virus Watch Prospective Cohort Study

**DOI:** 10.1101/2024.06.19.24309147

**Authors:** Sarah Beale, Alexei Yavlinsky, Gemma Moncunill, Wing Lam Erica Fong, Vincent Grigori Nguyen, Jana Kovar, Andrew C Hayward, Ibrahim Abubakar, Robert W Aldridge

## Abstract

**Background:** Early evidence suggests that people with Post-Covid Condition (PCC) may demonstrate aberrant immune responses post-infection; however, serological follow-up studies are currently limited. We aimed to compare SARS-CoV-2 serological responses to primary infection and vaccination in people who developed PCC versus those with an acute infection only.

**Methods:** Participants (*n*=2,010) were a sub-cohort of the Virus Watch community cohort study in England who experienced mild-moderate SARS-CoV-2 infections, completed surveys on persistent symptoms, and provided monthly finger-prick blood samples for serology. We compared the likelihood of post-infection seroconversion using logistic mixed models and the trajectories of anti-nucleocapsid (anti-N) and anti-spike (anti-S) antibodies using linear mixed models.

**Results:** Participants who developed PCC (*n*=394) had 1.8x the odds of post-infection seroconversion for anti-N antibodies compared to those with an acute infection only (*n*=1616) (adjusted odds ratio= 1.81 (95% confidence interval (CI) 1.16-2.90). Post-infection anti-N levels were persistently elevated in people with PCC (final log anti-N titres at 365 days 0.97, 95% CI 0.76-1.18) compared to those without (0.47, 95% CI 0.31-0.62). No differences were found in post-vaccination anti-S levels or trajectories before or after primary infection between participants with and without PCC; pre-vaccination anti-S responses could not be evaluated.

**Conclusion:** People with PCC demonstrated greater and more persistent anti-N antibody responses following primary infection compared to those with an acute infection only. Vaccination response pre- or post-infection did not systematically differ between groups. These findings extend emerging evidence around inflammatory and immune activation following infection in people with PCC.

## Background

While vaccination and effective treatments have reduced the threat from acute infection with Severe Acute Respiratory Syndrome Coronavirus 2 (SARS-CoV-2), ∼10-30% of people with mild-moderate infections and up to 50% of people with severe infections are left with chronic, interfering post-infectious symptoms (1,2). Post-Covid Condition (PCC) or Long Covid is a heterogeneous condition that can affect a range of organ systems, with commonly-reported symptoms including fatigue, dyspnoea, myalgia and/or arthralgia, and cognitive dysfunction (3). Understanding of the mechanisms underlying the development and maintenance of PCC is limited but evolving. Current evidence suggests multiple mutually non-exclusive mechanisms, including SARS-CoV-2 persistence and/or reactivation of other latent viruses, inflammatory pathologies and autoimmunity, and coagulopathies and endothelial dysfunction (1,4). The specific underlying mechanisms may vary by case and over time and interact with one another to produce the diverse range of symptoms reported (1,4). Aberrant immune response to SARS-CoV-2 may underlie the transition from acute infection to chronic illness, and longitudinal studies investigating this transition are required. Additionally, identifying immune correlates of PCC may provide useful clinical markers to assist with case identification as well as understanding aetiology.

While some individuals clear SARS-CoV-2 through their initial cellular and humoral innate immune response, the majority of people develop an adaptive immune response aimed at resolving the infection and leading to long-term immunological memory (5). Antibodies produced as part of this adaptive response include long-term Immunoglobulin G (IgG) antibodies targeting the SARS-CoV-2 spike and nucleocapsid structural proteins (5). The spike protein enables viral entry into host cells (6,7) and is the target of most licensed COVID-19 vaccines; consequently, anti-spike (anti-S) antibodies are produced in response to both natural infection and vaccination (8). The nucleocapsid protein is critical for the viral RNA genome packaging (6,7) and anti-nucleocapsid (anti-N) antibodies are produced in response to natural infection (9). The degree and persistence of individuals’ serological response to a SARS-CoV-2 immune challenge are influenced by a range of factors including age, sex, infection severity, comorbidities and immune-modulatory medications (10).

Early evidence suggests differences in the antibody response to infection in people who develop PCC compared to those who recover fully from SARS-CoV-2. Two-year post-infection follow-up of 31 people with PCC and 31 acute-only controls found that markers of inflammation and anti-S and anti-N antibodies were elevated in people with PCC, but normalised to the levels of the acute-only group after 12-24 months (11); participants were infected prior to the availability of COVID-19 vaccines, but later post-vaccination antibody responses appeared to be similar between groups. Similarly, a cohort of people with mild SARS-CoV-2 infections who were unvaccinated at the time of infection found that anti-S levels were higher at three months in women with PCC compared to those with acute infections only, and that levels of anti-N antibody were similar between both groups (12). A further cohort of 51 community cases followed-up twice for 5-6 months post-infection found persistently elevated anti-S and anti-N antibody levels in people with greater post-infectious symptomatology (13). The direction of findings in the literature was mixed, with a single-site hospital cohort of 107 patients finding persistently lower anti-N levels and similar levels of anti-S in people with PCC compared to those with an acute infection only; however, all participants in this cohort received antivirals likely limiting generalizability of findings (14). A further small, single-clinic study (*n*=61) monitoring anti-N levels over eight months post-infection found no difference between people with and without PCC (14,15).

Differences in post-infection antibody responses in people with PCC may be indicative of immune activation (11) or the persistence of SARS-CoV-2 virus or antigens during the transition from acute to chronic illness and in the maintenance of PCC symptoms, and warrants further investigation. Small sample sizes in the current literature, which is largely based on single-site cohorts, may introduce uncertainty into the comparison of people with PCC versus those with acute infections only. Furthermore, current studies are based exclusively on early-pandemic infections in unvaccinated people, and investigation including infections from later pandemic periods is warranted and would allow delineation of differences in post-vaccination and post-infection antibody responses in people with PCC.

### Aims and Research Questions

The primary aim of this study was to compare the anti-S and anti-N antibody dynamics related to primary SARS-CoV-2 infections in people who developed PCC and those with acute infections only. Participants were drawn from Virus Watch, a large UK community cohort including an adult sub-cohort who conducted monthly finger-prick serological sampling between February 2021 and March 2022. Research questions were:

1. Do odds of sero-conversion and anti-N antibody trajectories following infection differ between people who developed PCC and those with an acute infection only?
2. Do anti-S antibody trajectories differ following vaccination and/or infection between people who developed PCC and those with an acute infection only?

## Methods

### Ethics Approval and Consent

Virus Watch was approved by the Hampstead NHS Health Research Authority Ethics Committee: 20/HRA/2320, and conformed to the ethical standards set out in the Declaration of Helsinki. All participants provided informed consent for all aspects of the study.

### Participants

Participants (*n*=2,010) were a sub-cohort of Virus Watch (*n*=58,628), a household longitudinal cohort study of SARS-CoV-2 infections in England and Wales running since June 2020. Recruitment and methodology of the full cohort have been described in detail elsewhere (16,17).

Recruitment criteria into the Virus Watch study were residence in England or Wales, household size up to six people with consent or assent of all household members, access to an email address, and ability to complete English-language surveys. Households were recruited using several methods including SMS and postal recruitment and social media campaigns. Participants completed a detailed baseline questionnaire about demographic and clinical features for all household members, and subsequently completed weekly questionnaires about acute symptoms, SARS-CoV-2 tests, and vaccinations, and monthly questionnaires about detailed psychosocial and clinical topics tailored to the phase of the pandemic. A sub-cohort (*n*=19,555) of participants over 18 years of age and resident in England also completed monthly finger-prick antibody testing for SARS-CoV-2 antibodies (see Outcomes section below) with samples collected during the period between 24/02/2021-23/03/2022; participants in the current study were drawn from this sub-cohort.

Further inclusion criteria for the current study were:

1. returned at least one finger-prick antibody sample with a valid result for anti-S and/or anti-N antibodies,
2. completed survey(s) about new-onset long-term symptoms which covered symptom development between February 2020 and March 2023, and had binary classifiable PCC status (see Exposure section below),
3. had their first recorded SARS-CoV-2 infection detected via polymerase chain reaction (PCR) or lateral flow test (LFT) before end of serological follow-up,
4. mild-moderate infection (i.e., convalesced in the community without hospitalisation).

### Exposure

As we were interested in the difference between antibody titres and trajectories over time by PCC status, the primary exposures were binary PCC status and the interaction between PCC status and time since immunogenic events (i.e., infection and - where relevant - vaccination).

#### PCC Status

Participants were classified as having developed PCC if they reported one or more new-onset long-term symptoms following polymerase chain reaction (PCR) or lateral flow test (LFT) confirmed primary infection, with symptoms meeting the World Health Organisation consensus definition for PCC: interfering long-term symptom(s) which cannot be explained by another diagnosis with an onset within three months of SARS-CoV-2 infection and a duration of at least two months (18). Participants who had new-onset symptoms with a duration of less than two months or that developed outside of the three-month period following infection were excluded. Participants were classified as not having developed PCC if they completed all long-term symptom surveys and never reported new-onset symptoms at any point during follow-up. Complete follow-up was required for acute-only participants to minimise misclassification of undetected new-onset symptoms.

To determine PCC status, participants were sent a questionnaire about new-onset long-term symptoms as part of the Virus Watch monthly surveys. These surveys requested participants to indicate whether they had experienced any new-onset long-term symptoms and provide onset dates and duration of interfering symptoms (see (19) for further detail), which were used to determine PCC status according to the criteria above. The survey did not specify that these symptoms were linked to a SARS-CoV-2 infection, to avoid perceptions of PCC influencing participants’ answers. The survey was sent online to the Virus Watch cohort four times: in February 2021, May 2021, March 2022, and March 2023. Participants were asked to report new-onset long-term symptoms that developed within the previous year - covering the period between February 2020 and March 2023, which included the full period associated with antibody testing; only the May 2021 survey had a shorter recall period (from February 2020) as it was intended to supplement any non-response. While there was consequently some overlap in survey periods, symptoms could be matched by their onset date and consequently tracked if these overlapped across surveys.

#### Time Since Immunogenic Events

Time was defined as the number of days between an immunogenic event and the antibody test date, with the event varying depending on the outcome of interest (anti-N and anti-S). The date range for sample inclusion began at 0 (i.e. day of immunogenic event) and the upper limit was determined depending on the outcome, with anti-N models capped at 365 days while the anti-S models were capped at a maximum of 200 days due to stratification (see Statistical Analysis section below). For logistic models (see Statistical Analysis section), time since immunogenic event was categorised into the following bands previously used in Virus Watch research related to seroconversion to facilitate interpretation of odds ratios by time period (10): 0-29 days, 30-59 days, 60-89 days, 90-119 days, 120-269 days, 270+ days. In linear regression models, time in days was used to produce estimated antibody trajectories.

Infection was defined as evidence of first infection based on PCR or LFT based on linkage to UK national testing records or study-specific testing records. All participants had results available from linkage and also self-reported any SARS-CoV-2 tests taken across the study period in the weekly survey. PCR and LFT testing were also provided by the Virus Watch study during several periods, with the protocol varying over time (please see (19) for details).

COVID-19 vaccination status was determined based on linkage to UK national vaccination records as well as self-reported vaccinations collected as part of the weekly survey, and was coded as (0, 1, 2, 3 doses). Samples from participants who received additional doses - which were only available to a minority of the UK population (20) - were excluded after the third dose.

### Outcome

The outcomes of interest were anti-N and anti-S antibody titres, based on self-collected capillary blood samples (400-600 μl) collected between 24/02/2021-23/03/2022. Participants collected samples at home using test kits produced by the company Thriva and returned kits using prepaid priority postage. Serological testing was conducted in UK Accreditation Service accredited-laboratories using the Roche Elecsys Anti-SARS-CoV electrochemiluminescence assays targeting total immunoglobulin (predominantly IgG, but also IgA and IgM) to the nucleocapsid (N) protein and the receptor binding domain in the S1 subunit of the spike protein (21). Further details of the laboratory testing process for Virus Watch samples are provided elsewhere (10,22).

Antibody titres were expressed as semi-quantitative numeric values in form of cut-off indices (COIs) and log-transformed to base 10. For anti-N antibodies, the manufacturer-recommended seropositivity threshold was ≥1.0, with a sensitivity of 97.2-99.5% and specificity of 99.8% (23–25). Base-10 log transformed anti-N titres were included for samples taken between 0-365 days following PCR or LFT-confirmed primary infection. Samples that were collected following PCR- or LFT-confirmed reinfections were excluded due to the impact of reinfection on both antibody titres and unknown impact on long Covid symptomology; investigation into reinfections was beyond the scope of this analysis.

Samples that were anti-N seropositive within 5 days following infection or that demonstrated a four-fold rise in titres between sequential samples taken beyond 120 days following primary infection were also excluded to remove otherwise undetected reinfections, based on established timelines of conversion and trajectories (26).

For anti-S antibodies, the manufacturer recommended seropositivity threshold was ≥0.8, with a sensitivity of 97.9-98.8% and a specificity of 100% (23–25). Anti-S titres were subject to detection limits that changed over time to allow investigation into quantitative antibody levels in the highly vaccinated UK population, with limits changing from 250 u/mL between 24/02/2021 - 30/06/2021 (excluding a two-day pilot of the protocol change to increase detection limits), to 25000 u/mL between 01/07/2021 - 01/01/2022, and to 100000 u/mL between 01/01/2022 - 21/03/2022; the assay remained consistent and increased detection limits were obtained through dilution. Samples for anti-S were only included from 01/07/2021 due to a large number of samples reaching the low initial detection limit of 250 u/mL prior to this time point. The later change in the detection threshold was addressed through stratification (see Statistical Analysis section below). Samples were included if they occurred prior to primary infection (i.e., for vaccination only models) or following primary infection and prior to any confirmed reinfection. Samples following confirmed or suspected reinfection as described above were excluded for the remainder of follow-up. As with anti-N, anti-S titres were log-transformed to base 10.

### Stratification Variables

The following variables based on data collected in an online demographic survey upon study registration were used to test for effect modification and stratify models: sex at birth (male or female), and binary comorbidity status (presence of any condition on the UK NHS/government list denoting extreme clinical vulnerability or clinical vulnerability at COVID-19 (27). Please see ‘Conceptual Models and Effect Modification’ in the Statistical Analysis section for further details.

### Statistical Analysis

We used binary logistic mixed models to investigate how PCC status influenced the probability of seroconversion for anti-N. A random term was included to account for individuals submitting multiple samples. Separate models were constructed to evaluate probability of ever demonstrating anti-N seropositivity across the full follow-up period, as well as models evaluating anti-N seropositivity during the following time periods, to assess between-group differences in trajectories of seropositivity: 0-29 days, 30-59 days, 60-89 days, 90-119 days, 120-269 days, 270+ days. Results were expressed as odds ratios and predicted probabilities based on average marginal effects to facilitate between-group comparison over time. Seroconversion was investigated for anti-N only as this was the primary outcome and non-conversion is a more prominent feature of anti-N response (10); only 3 participants in the current study did not seroconvert for anti-S.

We used linear mixed models to investigate how PCC status influenced the trajectory of log anti-N and anti-S antibody titres. The exposure was the interaction between PCC status and time since immunologic event, and the outcome was log anti-N/anti-S antibody titres. Anti-N was modelled for all participants across 365 days of follow-up, and a sensitivity analysis was conducted including only samples from participants who seroconverted for N during the study period. As anti-S antibodies respond to both vaccination status and infection status and the combination of these events may differentially affect antibody titres, models were stratified according to these characteristics. Only samples following the increase of the cap to 25000u/ML were included, as a substantial number of samples with the early cap (250uML) reached this threshold, possibly precluding accurate estimations of titres and between-group differences. The sample period corresponded to periods of two-dose vaccination onwards in the Virus Watch study population, so anti-S models were constructed to investigate response to two-dose and three-dose vaccination as follows: pre-infection (i.e. vaccination response only), hybrid immunity with vaccination before the infection, and hybrid immunity with vaccination after the infection. A schematic diagram of these models illustrating the timing of immunogenic events is provided in Supplementary Figure 1. All anti-S models were capped at 200 days follow-up due to data availability and UK vaccination schedules, except for the post-second-dose anti-S model (infection most recent event), which was capped at 90 days due to low sample availability beyond this point.

We tested models with time modelled as a linear term, a quadratic term, and with a B-spline with a single knot at 120 days for anti-N (10), 30 days for post-second-dose anti-S models (28), and 14 days for post-third-dose anti-S models (29). Models were selected based on Bayesian Information Criterion values. The spline models were used for anti-N and for anti-S post-second dose (pre-infection and post-infection with vaccination as the most recent outcome); time was modelled using a linear term for the remaining anti-S models.

#### Conceptual Models and Effect Modification

Conceptual models underlying these analyses are presented in Supplementary Figure 2a for anti-N and Supplementary Figure 2b and 2c for anti-S antibodies. These analyses did not aim to estimate the causal effect of PCC status on antibody responses. Rather, differences in antibody responses to infection by PCC status were investigated to provide evidence for differential immune processes and/or viral persistence, which are proposed mechanisms for PCC development and which could not be directly measured here (denoted as node ‘U’ in Supplementary Figure 2a-c). Consequently, pre-infection demographic and clinical features and infection-related features (e.g. variant) are proposed to influence these unmeasured mechanisms, and are consequently conceptualised as effect modifiers rather than confounders. The direct effect of vaccination on anti-S antibody responses was addressed through stratification as described above. Infection severity was limited to mild-moderate community infections within this study based on inclusion criteria and study composition.

Pre-infection, vaccination-related anti-S responses were also investigated by PCC status (i.e., following later infection) to evaluate evidence for any pre-infection differences in immune response following challenge with the SARS-CoV-2 spike protein. The associated conceptual model is illustrated in Supplementary Figure 2c. As described for the infection-related models, demographic and clinical features are appropriately conceptualised as effect modifiers within this framework.

We consequently assessed effect modification by sex at birth and by comorbidity status for anti-N models covering the full study period. We included interaction terms adding sex and comorbidity status and evaluated evidence of the interaction providing additional explanatory power to the model using likelihood ratio tests. Associated results (i.e, predicted probabilities and predicted antibody trajectories) were presented stratified by sex and comorbidity status. These interaction tests were conducted for anti-N models only as anti-S models were already stratified according to vaccination status and samples were not sufficient to meaningfully assess three-way interaction for antibody trajectory across all models and timepoints. We lacked the sample size to assess effect modification for more granular variables, such as variant of infection.

## Results

Table 1 reports participants’ demographic characteristics overall and by PCC status. Of the 2010 included participants, 20% (*n*=394) developed PCC according to the WHO consensus definition following SARS-CoV-2 infection, while 80% (*n*=1616) experienced an acute infection only. The 2010 participants submitted 9466 included samples, with a median of 5 samples submitted per participant (interquartile range 3-6). Supplementary Figure 3 reports participant selection into the study. The number of samples within each regression model are reported overall and by PCC status in Supplementary Table 1; the majority of samples (79%, *n*= 7512) occurred prior to infection and were included in pre-infection, vaccination-related anti-S models only. Consequently, differences between participants’ vaccination status at the time of infection (Table 1) and the number of participants in post-vaccination, post-infection anti-S models (Supplementary Table 1) were due to lower sample availability post-infection. Table 1 reports on all participants, including those with only pre-infection samples available.

**Table 1.**
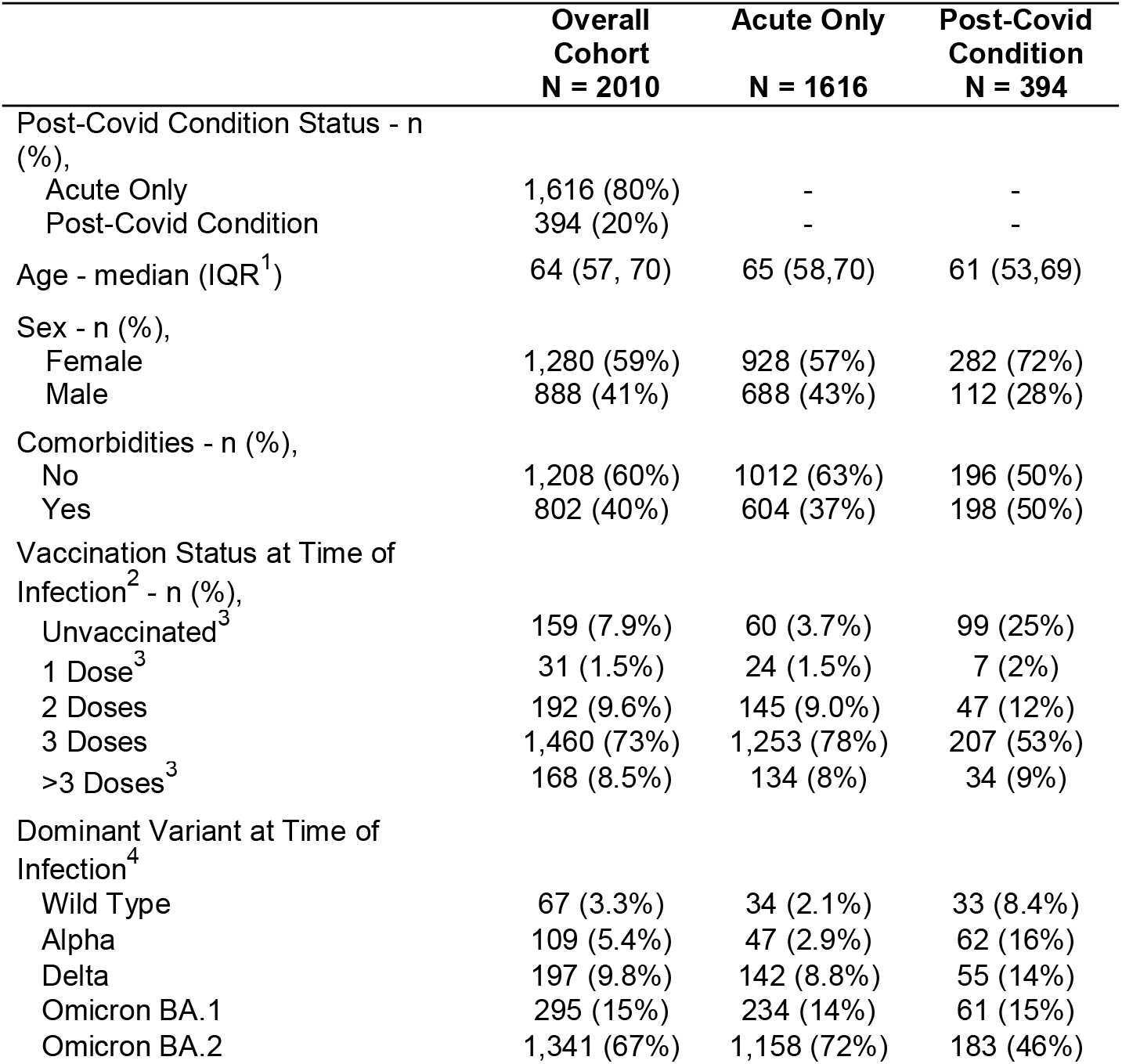
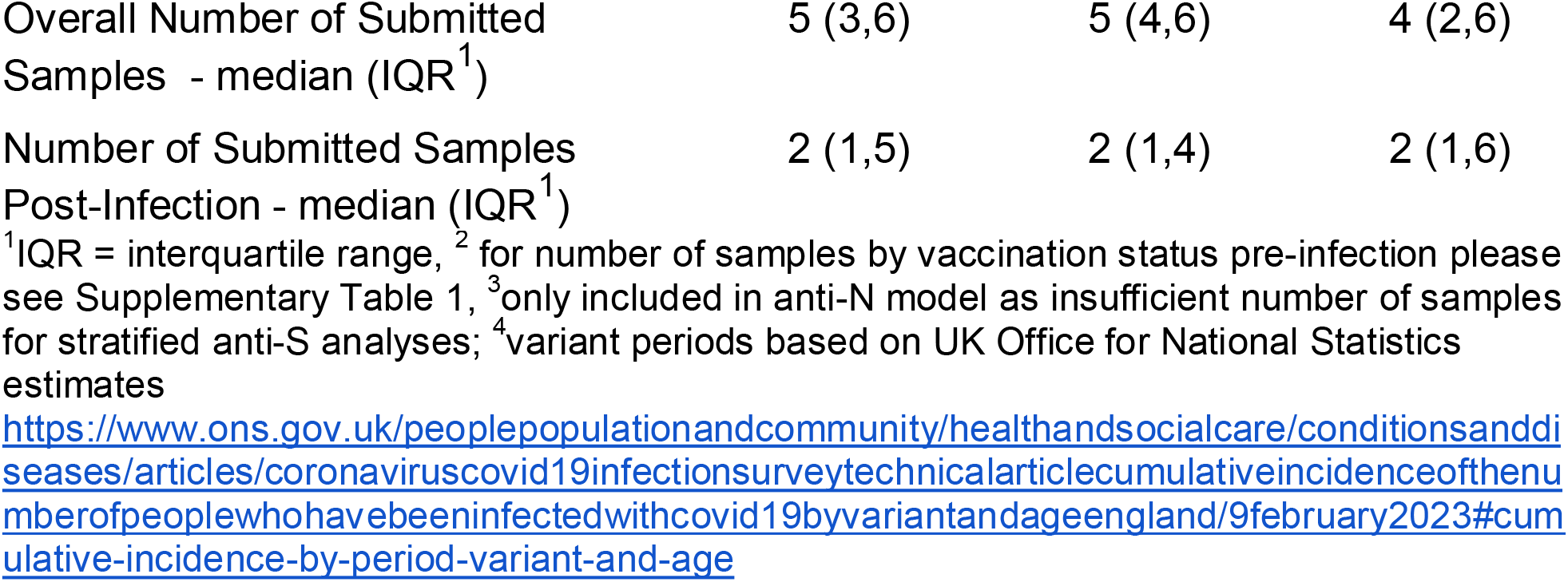
Participant Characteristics.

### Anti-Nucleocapsid Seropositivity by PCC Status

Across the full follow-up period, people with PCC were more likely to demonstrate detectable anti-N seropositivity after accounting for the effects of age, sex, and clinical vulnerability (OR= 1.81 (95% CI 1.16-2.90, *p*=0.01) compared to people with an acute infection only (see Supplementary Table 2 for odds ratios). Corresponding predicted probabilities (PP) for overall seroconversion were 84.7% (95% CI 78.9%-89.2%) amongst people with PCC versus 75.4% (95% CI 71.0% - 79.3%) for people with an acute infection only.

**Table 2.**
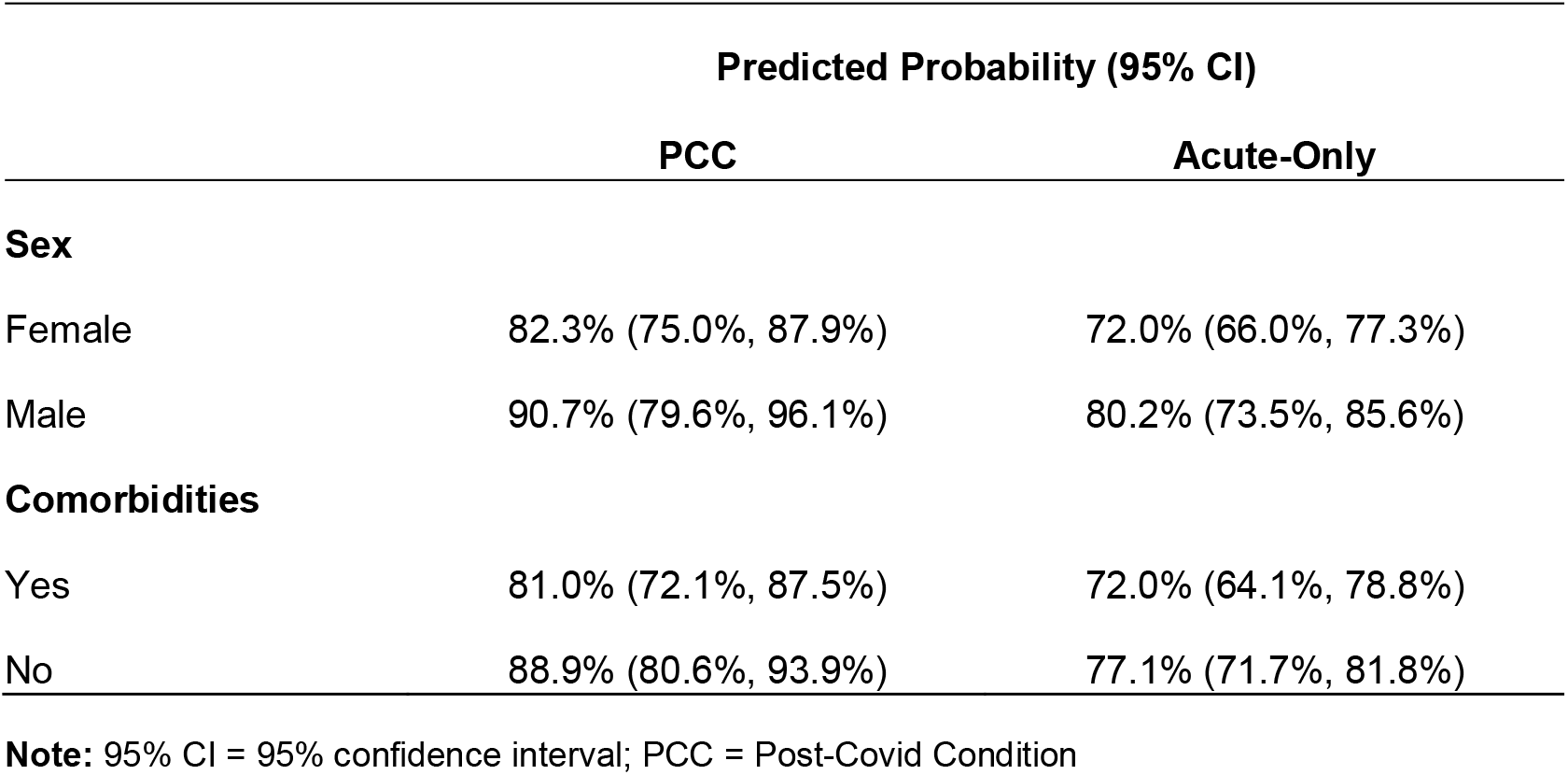
Predicted Probabilities of Anti-Nucleocapsid Seroconversion Stratified by Sex and Comorbidity Status (Overall Across Full Follow-Up Period)

When disaggregated by time since infection (see Figure 2 for predicted probabilities, and Supplementary Table 2 for odds ratios), people with PCC were more likely to demonstrate detectable anti-N seropositivity than people with an acute infection beyond the effect of chance during the following time periods: 60-89 days (OR=2.26, 95% CI 1.07-5.25, *p*=0.04), 90-119 days (OR=3.86, 95% CI 1.89-10.00, *p*=0.003), 120-269 days (OR=3.84, 95% CI 2.02-7.80, *p*<0.001), and 270+ days (OR=4.45, 95% CI 2.08-10.00, *p*<0.001). This corresponds with a pattern of increasing predicted probabilities of seropositivity during the initial periods following infection followed by a gradual decline in seropositivity in the acute-only group (78.3%, 95% CI 71.7-83.6% seropositive at 30-59 days versus 50.7%, 95% CI 39.1-62.3% seropositive at 270+ days) compared to persistently high seropositivity maintained across this period in the PCC group (85.2%, 95% CI 73.1-92.4% seropositive at 30-59 days and 82.1%, 95% CI 71.1-89.5% seropositive at 270+ days) (Figure 2).

**Figure 2.**
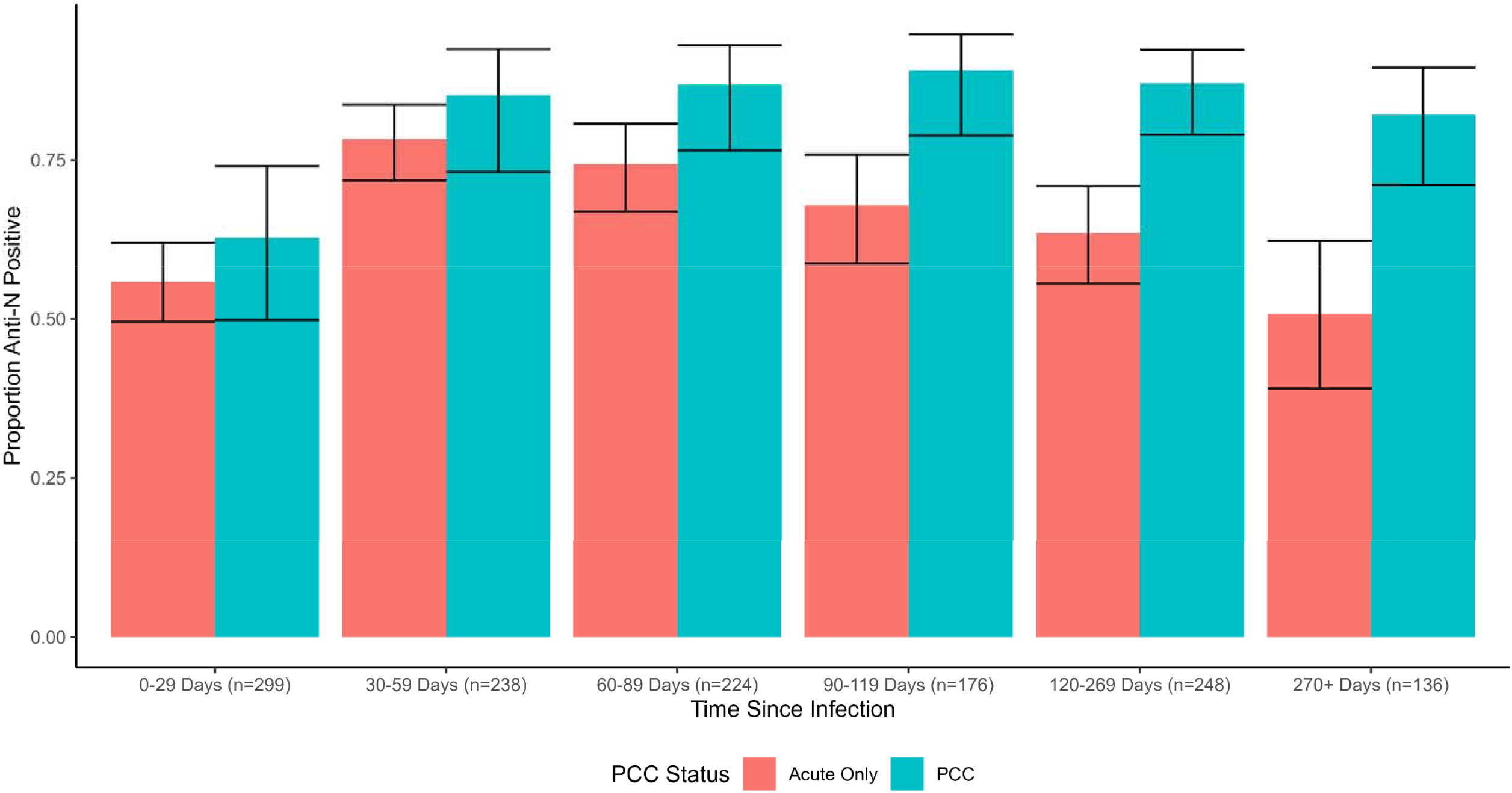
Predicted Probabilities of Detectable Anti-N Seropositivity in People with PCC versus Acute Infection Only, by Time Since Infection

#### Investigation into Effect Modification by Sex and Comorbidity Status

We found no evidence of an interaction between PCC status and sex or between PCC status and comorbidities beyond the effects of chance (Supplementary Table 3), indicating a similar effect of PCC status according to sex and comorbidity status. The corresponding stratified predicted probabilities of seroconversion (Table 2) demonstrate that in both male and female participants and in participants with and without comorbidities, people with PCC had a trend towards greater probability of seroconversion than those without; however, confidence intervals for predicted probability estimates overlapped for stratified models.

These investigations were included to investigate potential interactions between sex and comorbidity status and PCC status; for an appropriately powered and designed investigation of the main effects of sex on anti-N seropositivity, please see (10).

### Log Anti-N Levels over Time by PCC Status

Anti-N antibody trajectories are illustrated in Figure 3, with the solid lines illustrating regression estimates and the scatterplots illustrating raw trajectories. Individual regression coefficients are not interpretable in the presence of the interaction term and spline, and consequently results are presented graphically and quantified using estimated marginal predictions for key timepoints. Log antibody titres diverged by PCC status beyond the effect of chance from Day 24 post-infection, with an estimated log anti-N titre of 0.73 (95% CI 0.57-0.88) in people who developed PCC versus 0.46 (95% CI 0.36-0.56) in those with an acute infection only. Estimated titres were higher in the PCC group for the remainder of the follow-up period, following a similar trajectory in both groups with an initial exponential rise followed by a gradual waning. Titres peaked on Day 88 post-infection in the PCC group (estimated log anti-N titre=1.51, 95% CI 1.36-1.66) and on Day 81 in the acute-only group (predicted log anti-N titre=0.99, 95% CI 0.88-1.09). By 365 days post-infection, estimated titres had waned to 0.97 (95% CI 0.76-1.18) in the PCC group versus 0.47 (0.31-0.62) in the acute-only group.

**Figure 3.**
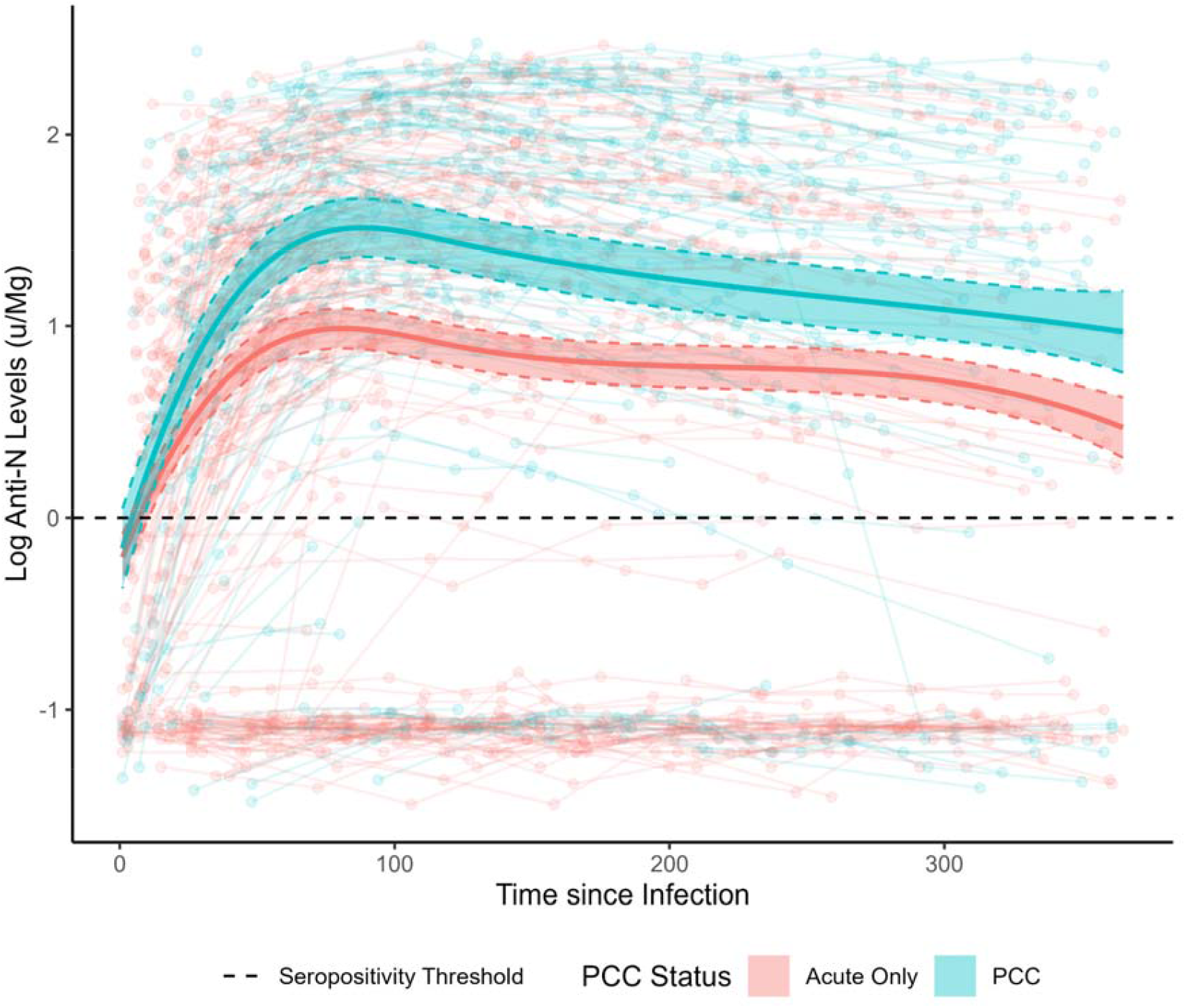
Anti-Nucleocapsid Antibody Trajectories in Participants with Post-COVID Condition versus Acute Infection Only **Note:** Anti-N= Anti-Nucleocapsid; PCC = Post-COVID Condition; solid lines indicate regression estimates for anti-N trajectories over time adjusted for age, sex and comorbidities; scatterplot indicates raw anti-N trajectories for individual participants; *n*=1094 samples from 604 participants

In the sensitivity analysis limited to participants who seroconverted for anti-N (Supplementary Figure 4), similar trajectories and between-group differences were identified by PCC status though differences were less pronounced and confidence intervals overlapped during later periods of follow-up beyond 200 days.

#### Investigation into Effect Modification by Sex and Comorbidity Status

Figure 4 presents predicted anti-N trajectories stratified by sex and comorbidity status, based on regressions including three-way interaction terms (see Statistical Methods). Likelihood ratio tests did not indicate that the interaction terms for sex (*p*=0.58) or comorbidity status (*p*=0.81) added explanatory value to the models. Similar trends were identified in the stratified models, with people with PCC demonstrating higher estimated antibody titres but a similar waning trajectory to those with an acute infection only. Confidence intervals for estimated titres overlapped between people with and without PCC in the subgroup who had comorbidities and - in the later part of follow up - in males. This reduced magnitude of difference by PCC status is likely to reflect sample size constraints - particularly during later periods of follow-up (illustrated by line chart in Figure 4) - but may reflect between group differences. Between-subgroup differences in titres according to sex and comorbidity status were in line with previous research and have been investigated directly elsewhere in Virus Watch using appropriate methodology (10).

**Figure 4.**
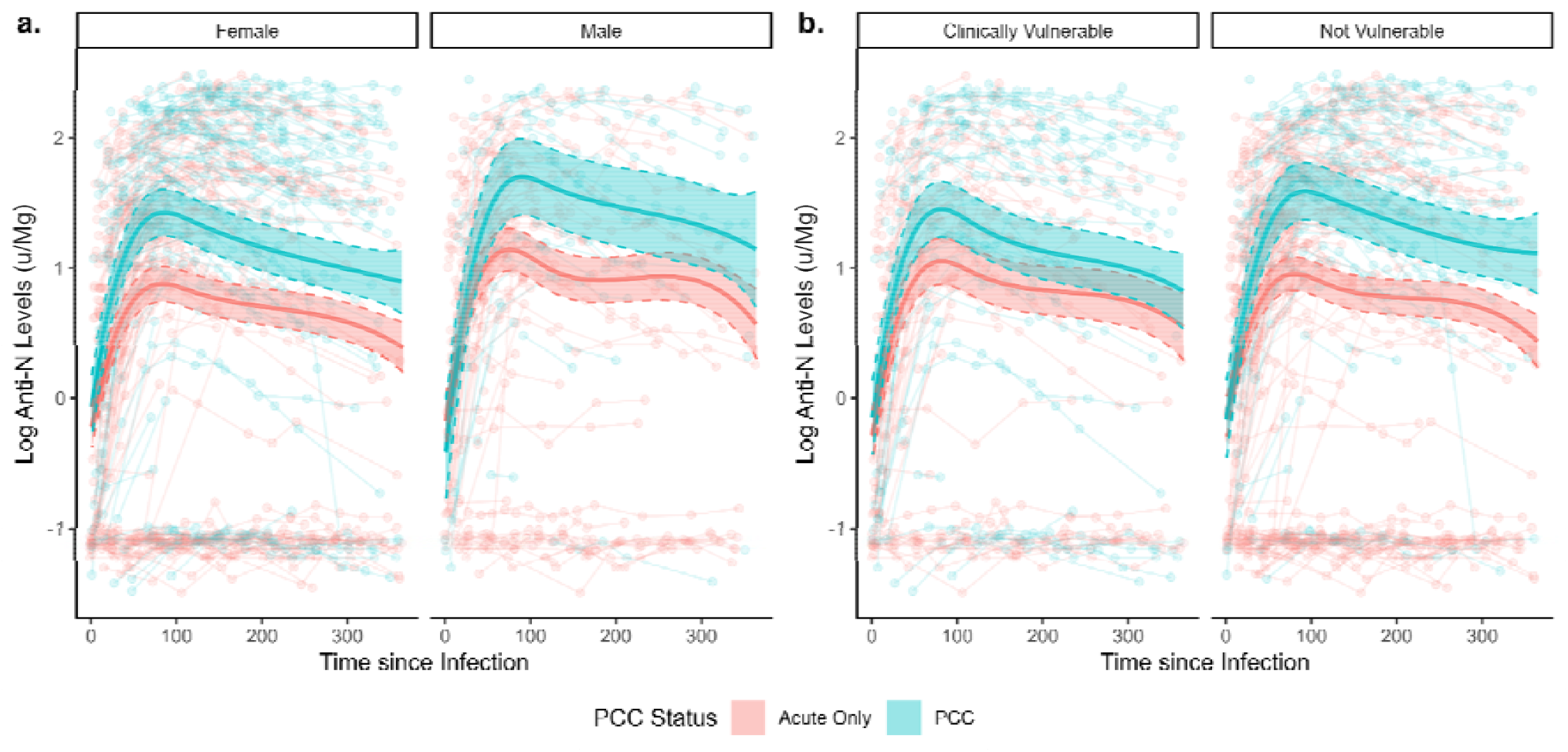
Anti-Nucleocapsid Antibody Trajectories in Participants with Post-COVID Condition versus Acute Infection Only, Stratified by Sex and Comorbidity Status

### Log Anti-S Levels over Time by PCC Status

Pre-infection anti-S responses to vaccination did not differ beyond the effects of chance in the 200 days following the second or third dose of COVID-19 vaccine (Figure 5), illustrated by overlapping estimated titres for both groups across the full follow-up period. Titres decreased over time following both doses, with an increase towards the end of post-second dose follow-up after log quadratic decline possibly reflecting unrecorded booster vaccination. There was a log-linear decline over time following the third dose. Similarly, there were no differences beyond the effect of chance in post-infection anti-S trajectories between participants with PCC versus those with an acute infection following two-dose or three-dose vaccination in any model (Supplementary Figure 5).

**Figure 5.**
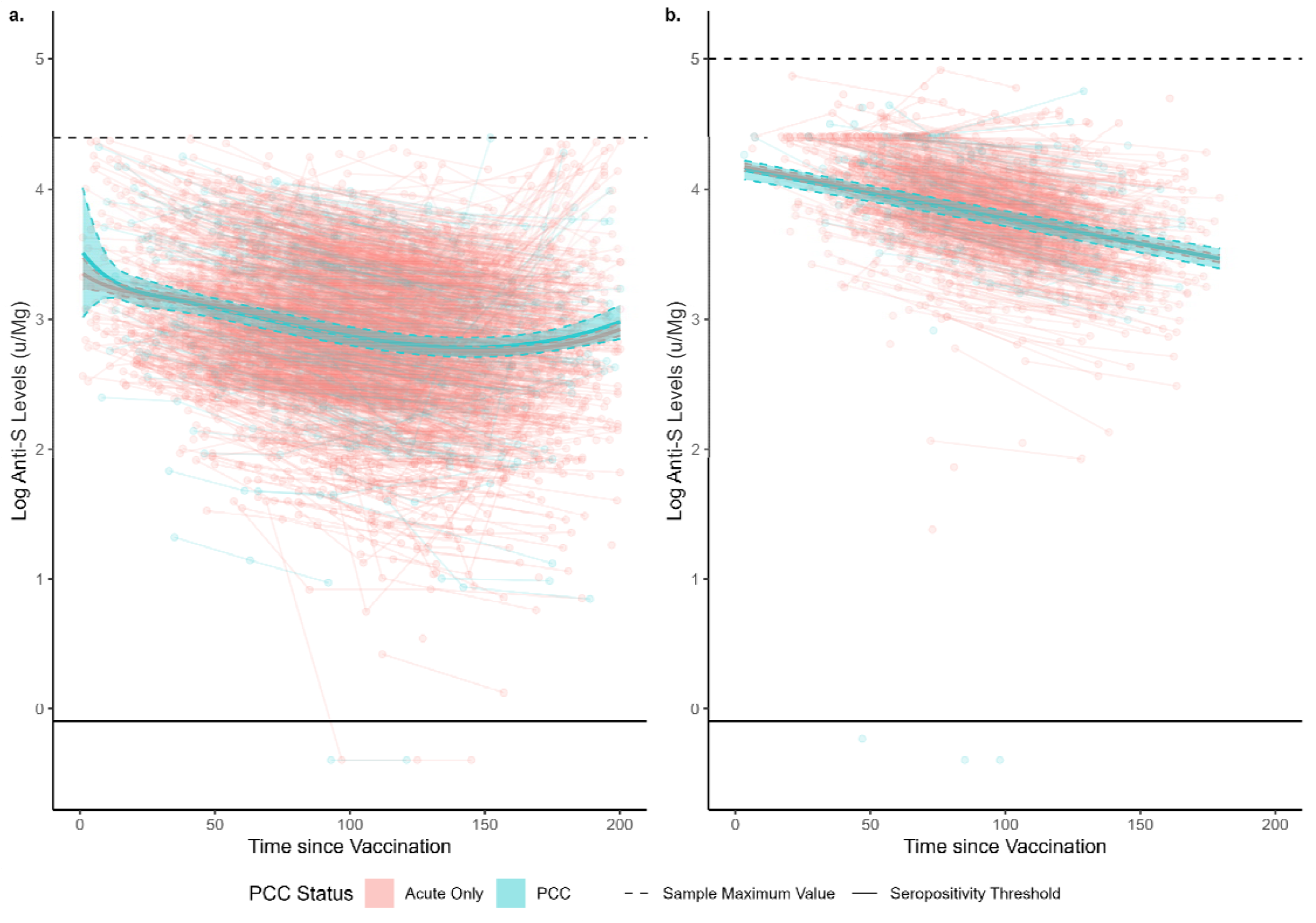
Vaccine-Induced (Pre-Infection) Anti-Spike Antibody Trajectories in Participants who Later Developed Post-COVID Condition versus Acute Infection Only following Two-Dose (a) and Three-Dose (b) Vaccination

## Discussion

We aimed to investigate differences in anti-N and anti-S antibody trajectories in people who developed PCC following a mild-moderate primary SARS-CoV-2 infection versus those with an acute infection only. Participants who developed PCC had 1.8x the odds of seroconversion for anti-N and demonstrated persistently higher levels of seropositivity compared to participants with an acute infection only, in whom levels of seropositivity decreased over time. Anti-N antibody titres diverged quickly following infection, with higher levels in participants who developed PCC than those with an acute infection only. While levels were elevated in PCC, anti-N trajectories were similar between the two groups and were characterised by an initial exponential increase followed by a gradual decline. Non-seroconversion in the acute-only group appeared to drive some of the difference in anti-N titres, although titres remained elevated - though less markedly - in the PCC group compared to the acute-only when limited to participants who seroconverted. Pre-infection vaccination responses and post-vaccination, post-infection anti-S titres did not differ according to PCC status.

Our findings broadly corroborate smaller community-based serological studies of early pandemic infections, which found evidence of stronger antibody responses in people with PCC compared to those with acute infections only (11,12),(13). The elevated, persistent anti-N response in people with PCC is suggestive of a stronger response to the acute infection, which could be driven by higher viral loads, a pro-inflammatory state and/or persistent viral antigens prompting the continued production of these antibodies (11). Notably, the current study enabled comparison of pre-infection anti-S responses to vaccination and found no evidence of a difference in levels or trajectory post-vaccination suggestive of pre-infection dysregulation in the PCC group. The findings of this study indicate areas for further investigation into the mechanisms underlying different infection-related serological responses in people with PCC and those without. Analysing isotypes (IgA, IgG) could inform how these responses may be stimulated. For example, antigen persistence in mucosal tissues may induce higher IgA levels (30). Similarly, analysing IgG subclasses may be informative, as they are linked to different effector functions (31,32) and the immunoglobulin glycosylation profiles, which are associated to inflammation and metabolic health (31). Given the evidence of elevated anti-N antibody levels in PCC, immunophenotyping B cells to investigate whether people with PCC demonstrate altered B cell phenotypes or expanded clonotypes that could be indicative of chronic infection or antigen persistence is warranted. More specific serological readouts may also enable the development of serological assessments indicative of PCC and/or help identify individuals at risk of PCC after acute infection.

Our findings are in line with those reported by Phetsouphanh et al. (11), who followed-up 31 people with PCC and 31 people without for two years, and who found elevated post-infection anti-S and anti-N antibody levels as part of a PCC immune phenotype suggestive of long-term innate and adaptive immune activation also including greater neutralising capacity and higher levels of S and N specific CD4+ T cells, PD-1, and TIM-3 expression on CD4+ and CD8+ T cells up to eight months post-infection. The length of follow-up in the current study was limited to 365 days, so it was not possible to determine whether antibody levels equalised in the groups beyond twelve months, as reported by Phetsouphanh et al. (11). The study authors suggested that vaccination and reinfection may have contributed to this later reduction in between-group variation in antibody titres, which may be reflected in the non-variation in post-infection anti-S titres in our highly vaccinated cohort.

The effect of PCC status on seroconversion for anti-N across the study period was consistent by sex and by comorbidity status, although the precision of results was likely affected by the relatively small subgroup sizes. Likewise, no statistical evidence of effect modification emerged between PCC status and sex or comorbidity status for anti-N antibody trajectories over time. However, confidence interval estimates overlapped amongst people with and without PCC who had comorbidities, and for males in the later part of follow-up; this may reflect sample size constraints but further stratified analyses in larger samples are recommended. Sex differences in the immune response in people with PCC have been previously characterised for other immune parameters (33) and are plausible according to comorbidities. However, the findings around effect modification in this study specifically concerned comparison of antibody trajectories between people with and without PCC over time, rather than direct comparison of antibody levels and trajectories amongst people with PCC by sex and comorbidity status, which was beyond the scope of this analysis. Investigation into more granular potential effect modifiers - such as variant of infection - were impeded by sample availability.

### Strengths and Limitations

Strengths of this study included the community-based sample drawn from across England, which was, to our knowledge, the largest PCC-related serological follow-up study to date. PCC was classified according to the WHO consensus definition. The cohort spanned infections beyond the first pandemic wave and enabled delineating the response to infection and vaccination as well as adjustment for demographic and clinical factors. Self-collection of serological samples enabled monthly sampling which provided more granular data around antibody trajectories than available in studies with less frequent follow-up.

While the overall sample size of this study was large in the context of serological PCC research, subgroup sizes were relatively small over time - particularly at later follow-up time points - and it was not possible to conduct analyses stratified by demographic and clinical characteristics, notably specific clinical conditions that may affect the immune response, variant of infection, and features/phenotypes of PCC. Phenotypes of PCC were beyond the scope of this study and relevant analyses are planned within the cohort. Other key limitations included the early threshold on anti-S testing precluding comparison of anti-S antibody trajectories in unvaccinated participants and those who had received a single dose of vaccination. This study focused on primary infections. However, some infections may have been misclassified reinfections due to the lack of testing early in the pandemic; detection of later reinfections was supported by access to swab testing and detection through serology in this cohort though undetected reinfections remain possible. This study was based on mild community cases and may not generalise to more severe cases or people who received specific COVID-19 treatments. While age, sex, and comorbidities were accounted for in regressions and the average marginal effect estimates produced from these models, the sample was not representative of the English population. The sub-cohort of participants who self-selected to complete the PCC survey and who provided serological samples may be particularly motivated regarding pandemic-related health behaviours including testing and symptom monitoring, or motivated by long-term symptomatology; while this may have impacted the composition of the sample and prevalence of people with PCC, its impact on antibody trajectories is likely minimal. The follow-up period was limited to 365 days for anti-N and shorter periods for anti-S due to model stratification, and longer follow-up with monitoring of PCC symptomatology is recommended.

## Conclusions

We identified a greater likelihood of seroconversion and elevated anti-N antibody levels in the 365 days following infection in people with PCC compared to those with an acute infection only based on a sample of community SARS-CoV-2 cases in England. These findings build upon previous evidence around altered adaptive immune responses in PCC suggestive of chronic post-infection inflammation and immune activation and/or viral persistence in people with PCC. Pre-infection responses to vaccination did not differ between people who later developed PCC following primary infection and those who recovered fully from their primary infection. Further direct investigation into immune processes following infection in people with PCC, as well as its relationship with symptom presentation, persistence, and severity is recommended. Building upon this evidence, further understanding of the immune processes underlying PCC is required to inform the development and selection of evidence-based treatments for the condition.

## Data Availability

We aim to share aggregate data from this project on our website and via a 'Findings so far' section on our website https://ucl-virus-watch.net/. We also share some individual record level data on the Office of National Statistics Secure Research Service. In sharing the data we will work within the principles set out in the UKRI Guidance on best practice in the management of research data. Access to use of the data whilst research is being conducted will be managed by the Chief Investigators (AH and RA) in accordance with the principles set out in the UKRI guidance on best practice in the management of research data. We will put analysis code on publicly available repositories to enable their reuse.

## Data Availability

We aim to share aggregate data from this project on our website and via a “Findings so far” section on our website—https://ucl-virus-watch.net/. We also share some individual record level data on the Office of National Statistics Secure Research Service. In sharing the data we will work within the principles set out in the UKRI Guidance on best practice in the management of research data. Access to use of the data whilst research is being conducted will be managed by the Chief Investigators (AH and RA) in accordance with the principles set out in the UKRI guidance on best practice in the management of research data. We will put analysis code on publicly available repositories to enable their reuse.

