## Supplementary Materials for "Anti-Nucleocapsid and Anti-Spike Antibody Trajectories in People with Post-Covid Condition versus Acute-Only Infections: Results from the Virus Watch Prospective Cohort Study"

**Supplementary Figure 1.** Schematic Diagram of Antibody Trajectory Models

a. **Anti-Nucleocapsid Model**

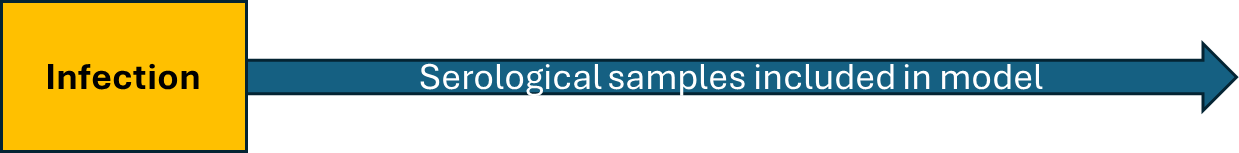

b. **Anti-Spike Models** (Pre-Infection Vaccination-Only and Post-Infection Hybrid Immunity Models)

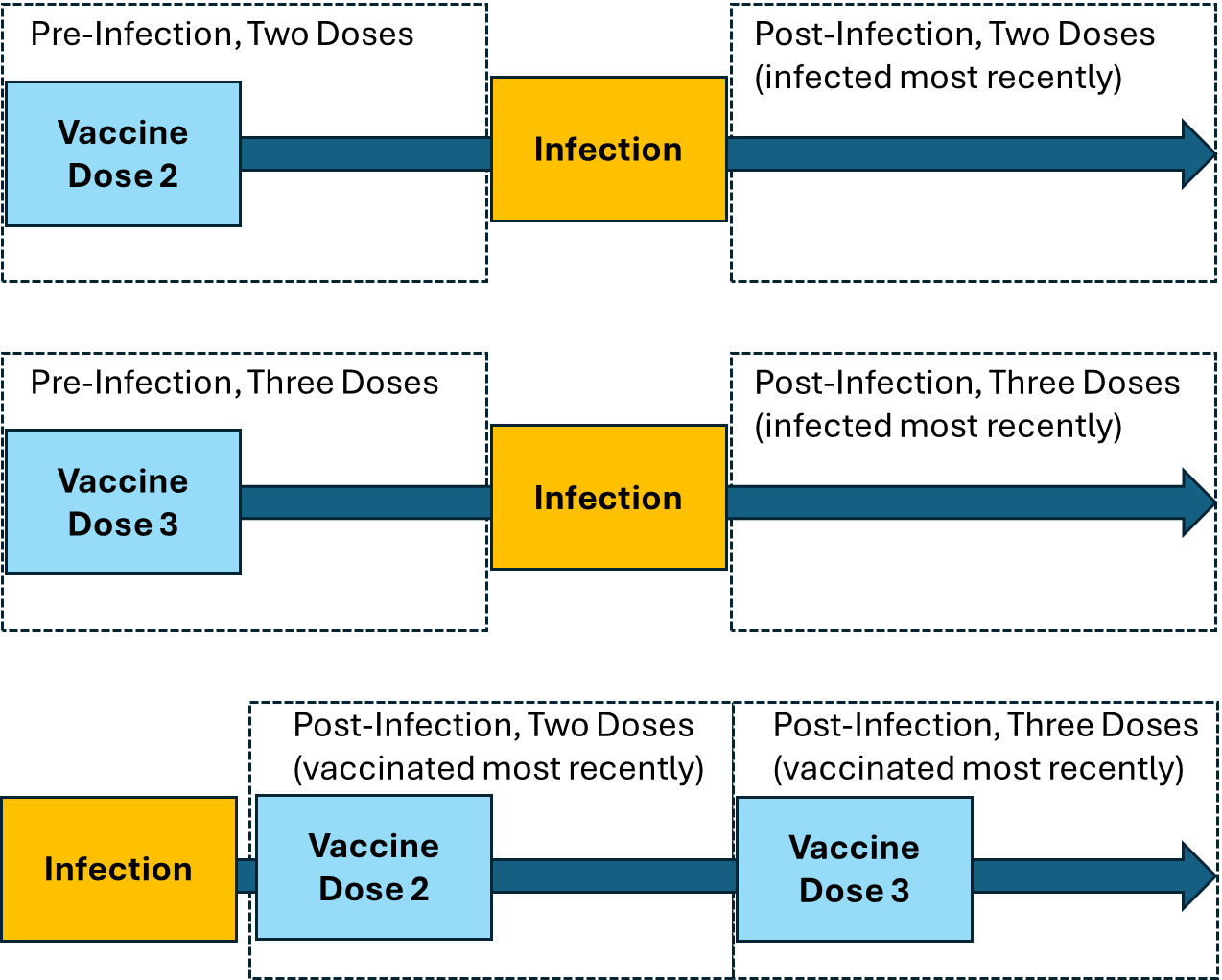

**Note:** The anti-Spike diagram above intends to provide examples of the timing of relevant immunogenic events (vaccination and/or infection) within each model. The distance between these events can vary, and events may be combined beyond the illustration above, which was simplified for clarity (e.g., dose two of vaccination may occur prior to infection and dose three afterwards, etc.)

**Supplementary Figure 2a.** Conceptual Model of Analysis: Post-Infection Anti-Nucleocapsid Antibody Response

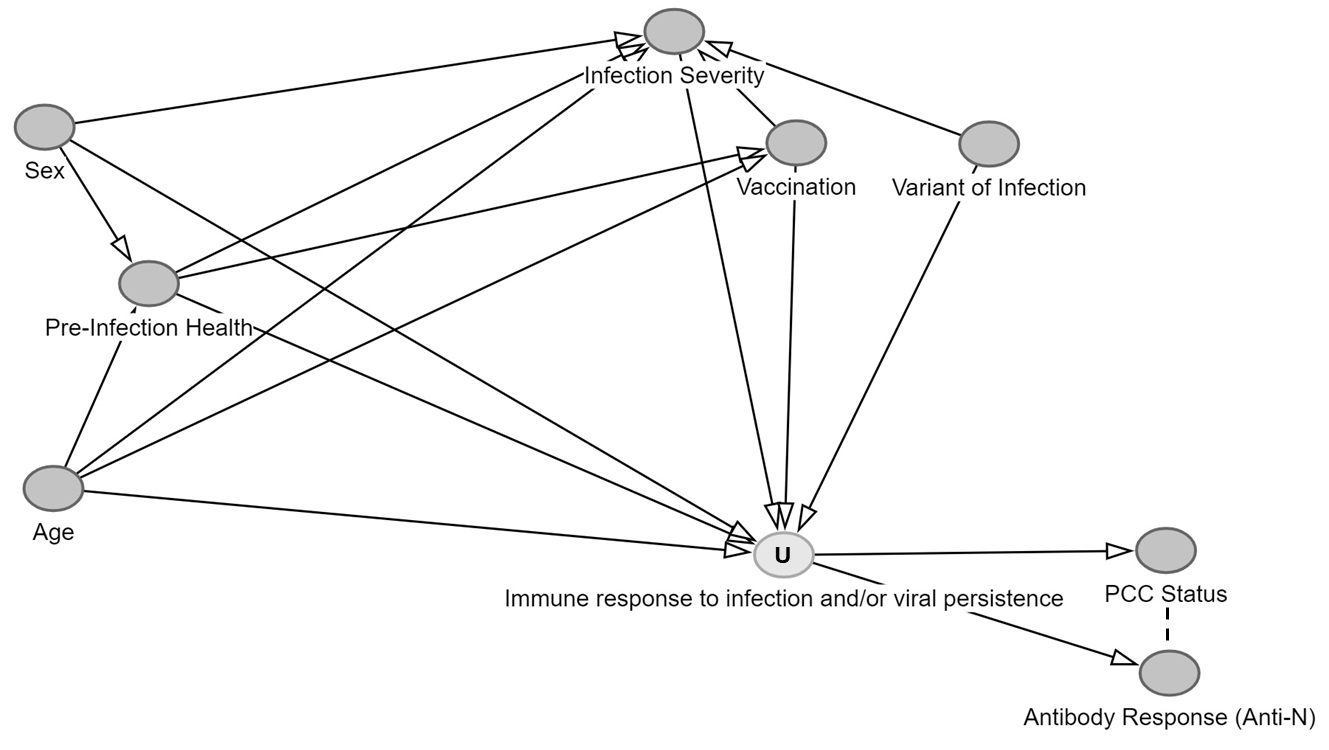

**Supplementary Figure 2b.** Conceptual Model of Analysis: Post-Infection Anti-Spike Antibody Response

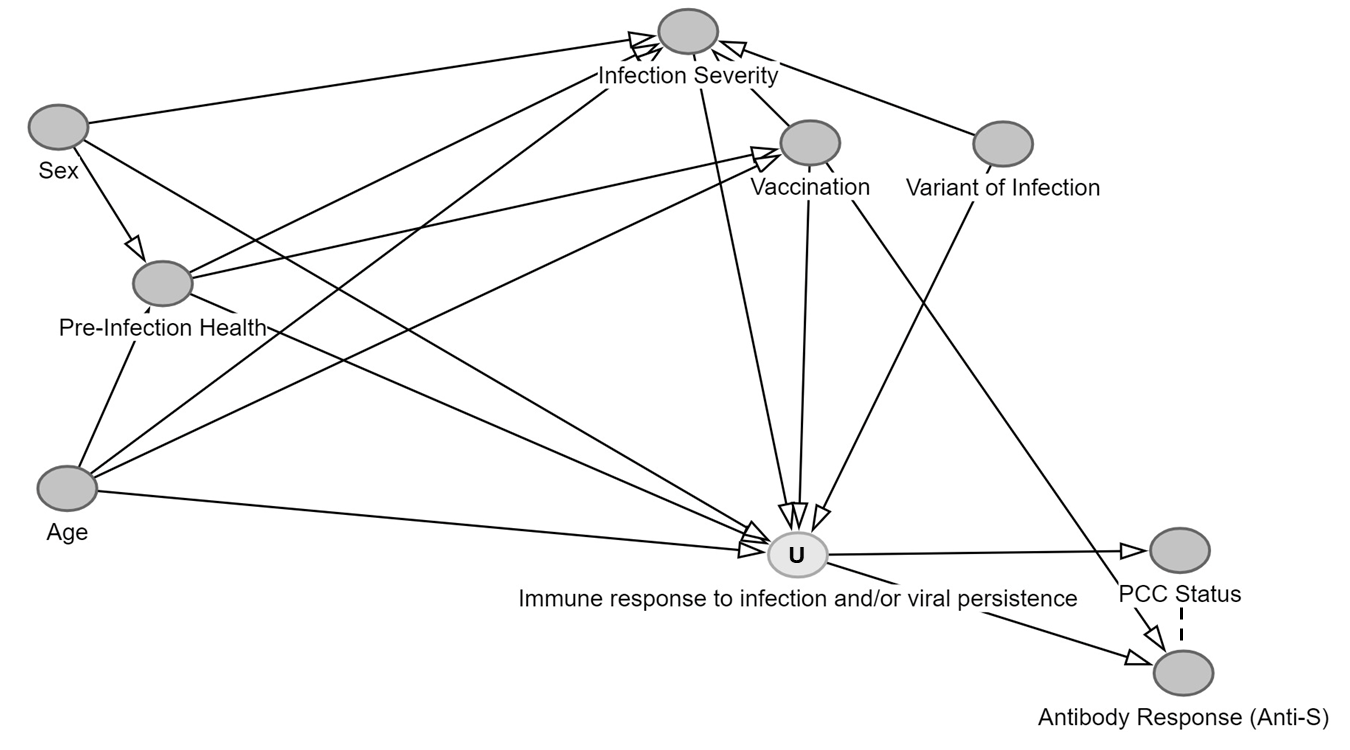

**Supplementary Figure 2c.** Conceptual Model of Analysis: Post-Vaccination (Pre-Infection) Anti-Spike Antibody Response

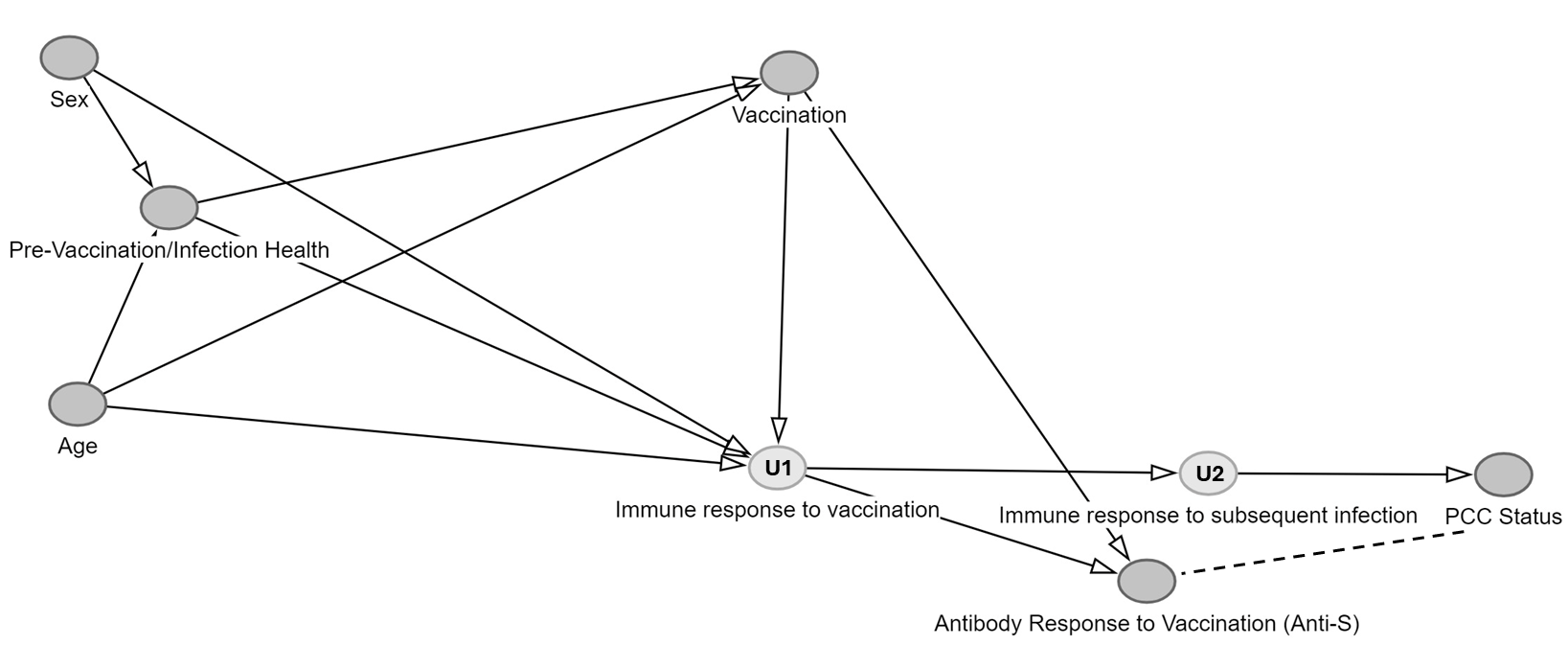

**Supplementary Figure 3.** Flowchart of Participant Selection

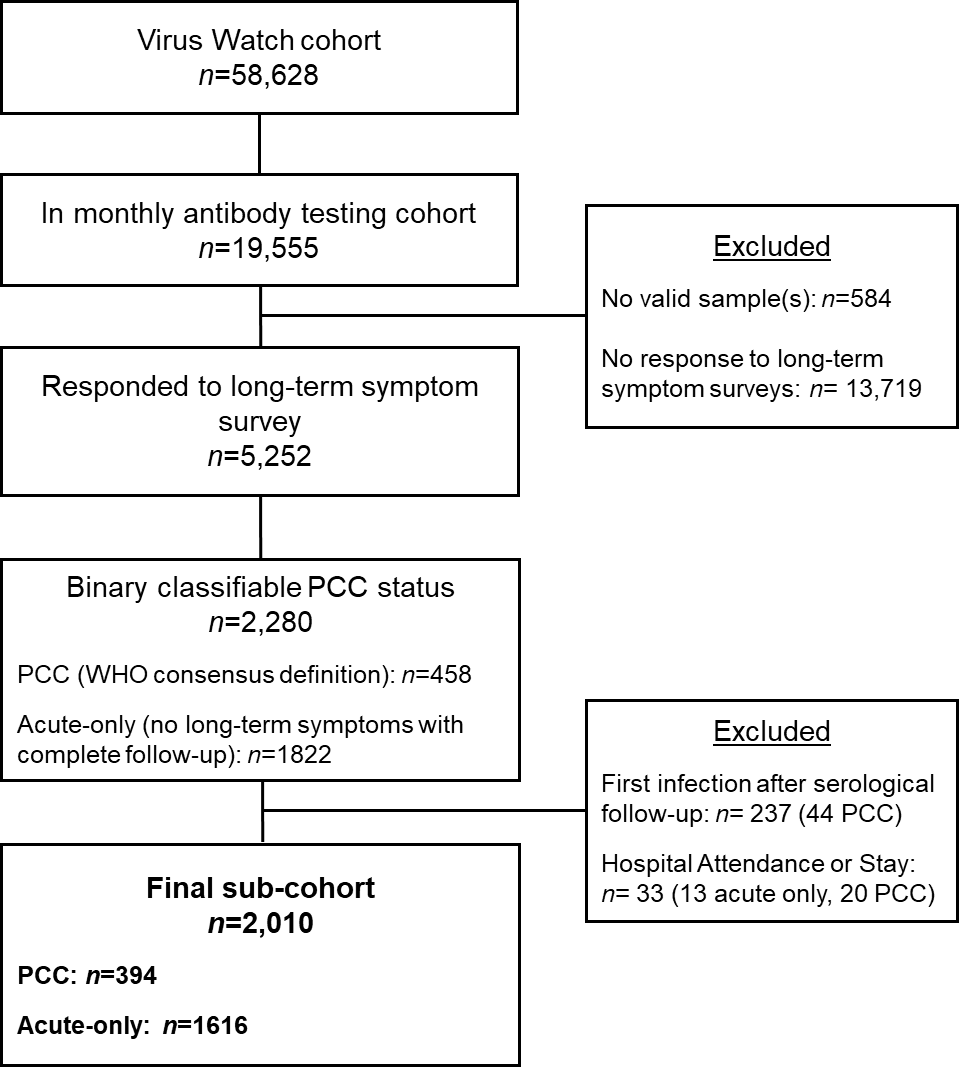

**Note:** PCC=Post-Covid Condition

**Supplementary Table 1.** Number of Samples per Antibody Trajectory Model

| **Model** | **Overall (*n*)**** | **Acute only (*n*)** | **PCC (*n*)** |
| --- | --- | --- | --- |
| Anti-N | 1899 | 1229 | 670 |
| Anti-N (seropositivity sensitivity analysis) | 1411 | 835 | 576 |
| Anti-S (pre-infection, two doses) | 5461 | 4775 | 686 |
| Anti-S (pre-infection, three doses) | 2051 | 1790 | 261 |
| Anti-S (post-infection, two doses - vaccinated most recently) | 455 | 241 | 214 |
| Anti-S (post-infection, two doses - infected most recently) | 93 | 73 | 20 |
| Anti-S (post-infection, three doses - vaccinated most recently) | 356 | 299 | 57 |
| Anti-S (post-infection, three doses - infected most recently) | 186 | 133 | 53 |

PCC=Post-Covid Condition

**Note: the sum of samples across models exceeds the total number of samples in the study (*n*=9466) due to testing for both S and N antibodies leading to inclusion of samples in multiple models.

**Supplementary Table 2.** Frequencies and Odds of Detectable Anti-N Seropositivity in People with PCC versus Acute Infection Only, by Time Since Infection

| **Group** | **Total (Acute Only)** | **Seropositive (Acute Only)** | **Total (PCC)** | **Seropositive (PCC)** | **OR** | **95% CI** | **p-value** |
| --- | --- | --- | --- | --- | --- | --- | --- |
| Overall | 410 | 309 (75%) | 190 | 161 (85%) | 1.81 | 1.16, 2.90 | 0.010 |
| 0-29 Days | 240 | 134 (56%) | 59 | 37 (63%) | 1.33 | 0.75, 2.42 | 0.3 |
| 30-59 Days | 184 | 144 (78%) | 54 | 46 (85%) | 1.60 | 0.73, 3.90 | 0.3 |
| 60-89 Days | 156 | 116 (74%) | 68 | 59 (87%) | 2.26 | 1.07, 5.25 | 0.043 |
| 90-119 Days | 112 | 76 (68%) | 64 | 57 (89%) | 3.86 | 1.69, 10.0 | 0.003 |
| 120-269 Days | 148 | 94 (64%) | 100 | 87 (87%) | 3.84 | 2.02, 7.80 | <0.001 |
| 270+ Days | 69 | 35 (51%) | 67 | 55 (82%) | 4.45 | 2.08, 10.0 | <0.001 |
| PCC=Post-Covid Condition; OR = odds ratio; CI = confidence interval | | | | | | | |

**Supplementary Table 3.** Odds Ratios and Likelihood Ratio Tests for Interaction between Sex and Comorbidity Status and PCC Status on Anti-N Seropositivity (Overall Model Across Full Follow-Up Period)

**Note:** In the presence of an interaction term, main effects cannot be interpreted directly. For the main effect of PCC status, please see Supplementary Table 2 and Figure 2. Main effects of sex and comorbidities on anti-nucleocapsid seroconversion have been investigated elsewhere in the Virus Watch data^1^ using appropriate methodology and samples.

| **Group** | **Characteristic** | **OR**^1^ | **95% CI** | ***p*-value** | **LRT *p*** |
| --- | --- | --- | --- | --- | --- |
| Sex | PCC Status (see note above) | 1.81 | 1.09, 3.10 | 0.026 |  |
|  | Sex (see note above) | 1.58 | 0.99, 2.55 | 0.059 |  |
|  | **PCC Status * Sex** | **1.33** | **0.46, 4.50** | **0.6** | **0.05** |
| Comorbidities | PCC Status (see note above) | 2.37 | 1.20, 5.12 | 0.018 |  |
|  | Sex (see note above) | 0.76 | 0.48, 1.22 | 0.3 |  |
|  | **PCC Status * Sex** | **0.70** | **0.26, 1.77** | **0.5** | **0.16** |

**Note:** OR = odds ratio; 95% CI = 95% confidence interval; LRT = likelihood ratio test; PCC = Post-Covid Condition

^1^ Navaratnam AM, Shrotri M, Nguyen V, Braithwaite I, Beale S, Byrne TE, Fong WL, Fragaszy E, Geismar C, Hoskins S, Kovar J. Nucleocapsid and spike antibody responses following virologically confirmed SARS-CoV-2 infection: an observational analysis in the Virus Watch community cohort. International Journal of Infectious Diseases. 2022 Oct 1;123:104-11.

**Supplementary Figure 4.** Anti-Nucleocapsid Antibody Trajectories in Participants with Post-COVID Condition versus Acute Infection Only: Sensitivity analysis limited to anti-N seroconverters

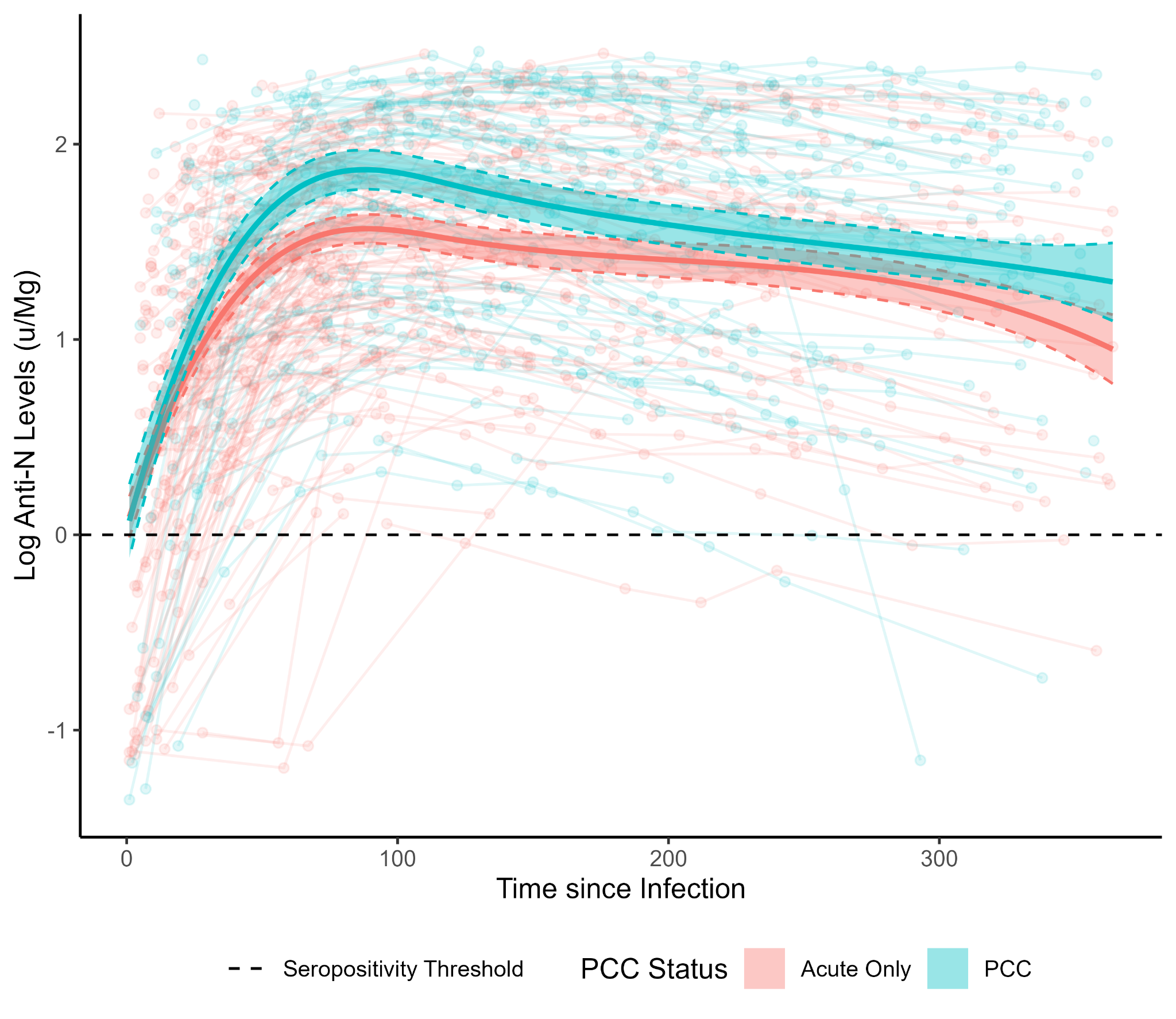

**Note:** *n*=1422 samples from 475 participants within this sensitivity analysis

**Supplementary Figure 5.** Post-Infection, Post-Vaccination Anti-Spike Antibody Trajectories in Participants with Post-COVID Condition versus Acute Infection Only: Post-two-dose vaccination with vaccination (a) and infection (b) most recently and and three-dose vaccination with vaccination (a) and infection (b) most recently

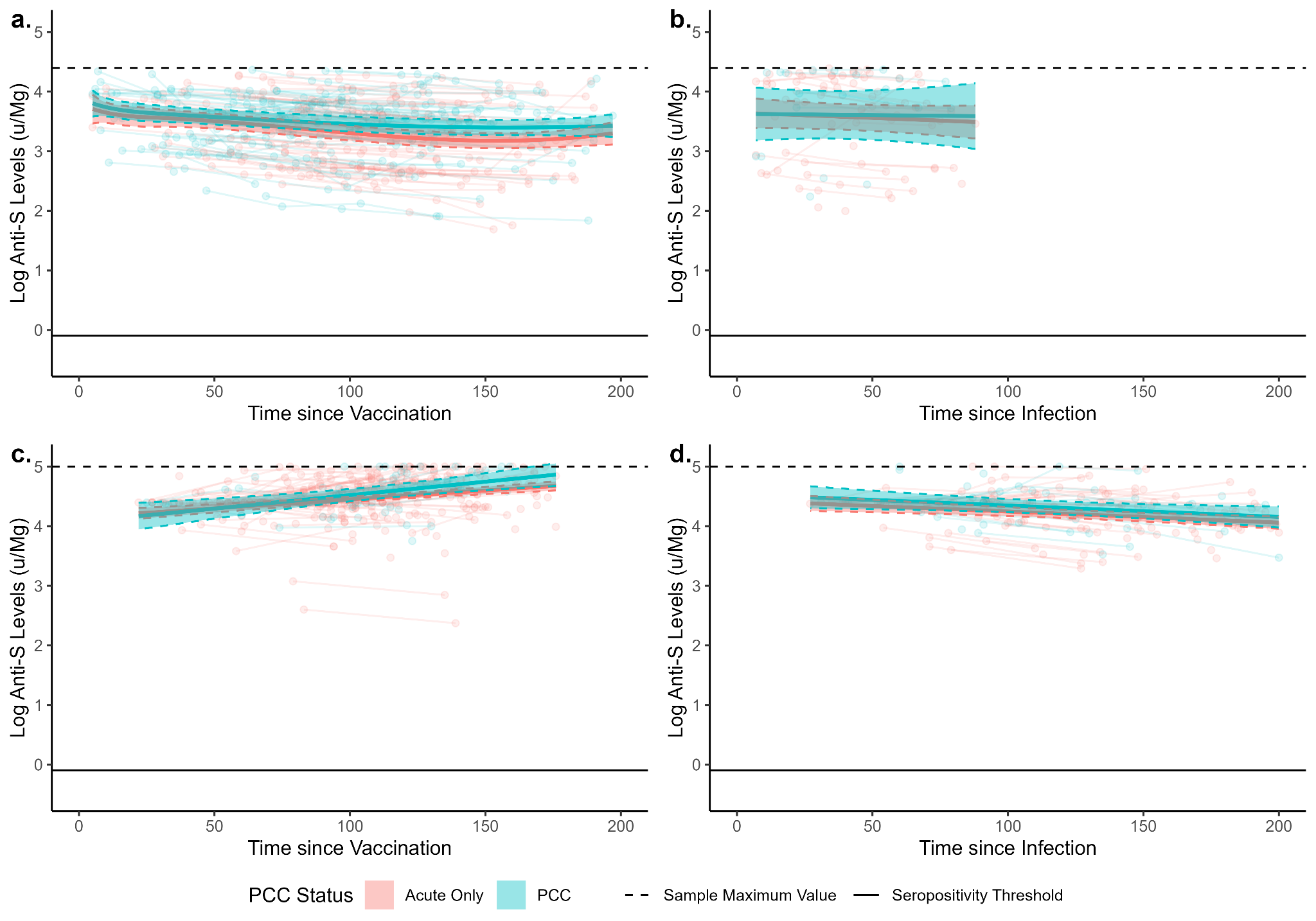
